# From Thought to Speech: Integrating a Low-Cost Electroencephalography Device with AI to Decode Neural Language Signals in Amyotrophic Lateral Sclerosis Patients

**DOI:** 10.64898/2026.01.18.26344355

**Authors:** Shaumprovo Debnath, Cassie N. Chan, Randee Kent, Prasanna Kolar, Eithan Kotkowski

**Affiliations:** Keystone School, 119 E Craig Pl, San Antonio, TX 78212, USA; Department of Medicine, University of Texas Health San Antonio, 7703 Floyd Curl Dr., San Antonio, TX 78229, USA; Department of Neurology, University of Texas Health San Antonio, 7703 Floyd Curl Dr., San Antonio, TX 78229, USA; Southwest Research Institute, 6220 Culebra Road, San Antonio, TX 78238, USA

**Keywords:** amyotrophic lateral sclerosis, dysarthria, brain-computer interface, machine learning, convolutional neural network

## Abstract

**Purpose:** Nearly all amyotrophic lateral sclerosis (ALS) patients develop dysarthria, with many progressing to anarthria and global expressive communication failure despite preserved consciousness. Despite the severity of this communication loss, available augmentative communication technologies remain limited. Brain-computer interface (BCI) technology provides a theoretically compelling approach for decoding speech directly from neural activity. Current BCI technologies are predominantly invasive, relying on surgically implanted microelectrode arrays that constrain feasibility and clinical accessibility. In this proof-of-concept study, we evaluated the feasibility of using a low-cost, noninvasive EEG device to record brain activity during subvocalized word production in patients with ALS and to decode these signals back into word-level outputs using a machine learning approach.

**Patients and methods:** Data were collected from five patients with ALS during subvocalized the words YES, NO, HELP, SUN, and WATER. Data from three participants met inclusion criteria and were analyzed for this report. Data from two participants were excluded due to movement-related artifacts. EEG signals were preprocessed using a Butterworth bandpass filter, after which the dataset was partitioned into training, validation, and testing subsets. A convolutional neural network (CNN) was then trained to decode the EEG features back into their corresponding subvocalized word labels.

**Results:** A total of 3,819 samples were analyzed, with each sample corresponding to EEG data from a single subvocalized word. Median classification accuracies on the held-out test set for the three ALS participants were 40.17%, 32.95%, and 54.76%, respectively. All accuracies were significantly greater than chance-level performance (20%) under the null hypothesis (p <0.001).

**Conclusion:** These results demonstrate that EEG signal patterns associated with subvocalized words can be detected and decoded using CNN. Collectively, the findings support the feasibility of a non-invasive EEG-based brain-computer interface as a potential communication modality for individuals with ALS and patients with severe physiological speech impairment.

## Introduction

Amyotrophic lateral sclerosis (ALS) is characterized by progressive degeneration of upper and lower motor neurons, leading to loss of voluntary motor control. Nearly all patients develop dysarthria, and many ultimately progress to anarthria with profound expressive communication failure despite preserved consciousness. As bulbar and limb motor pathways degenerate, conventional augmentative and alternative communication strategies – such as eye-tracking or switch-based systems – become increasingly burdensome and less effective.

Brain-computer interface (BCI) technology enables communication between the brain and an external device by acquiring, processing, and translating neural signals into executable commands. BCI technologies offer a means of bypassing impaired neuromuscular pathways by translating neural activity directly into external commands.^1,2^ To date, however, BCI studies in ALS patients have been limited in number and have relied on predominantly invasive approaches involving surgically implanted microelectrode arrays or cortical prostheses. Employing BCI technologies prior studies reported communication via typing interfaces,^3^ auditory neurofeedback for word formation,^4^ or direct decoding of cortical activity.^5^ These studies also required resource-intensive neuroimaging-guided systems such as functional magnetic resonance imaging for precise electrode localization, substantially increasing procedural complexity, cost, and barriers to clinical adoption.

To address these limitations, in this proof-of-concept study we evaluated the feasibility of using a low-cost, noninvasive electroencephalography (EEG) device combined with a machine learning framework to decode brain electrical signals generated during subvocalized word production in individuals with ALS. By targeting cortical speech-planning activity rather than overt motor output, this approach sought to bypass degenerating bulbar motor pathways and provide a potential communication modality for ALS patients with severe expressive impairment.

## Materials and Methods

Between July 10 and September 24, 2024, medical records of 122 patients with ALS registered at the University of Texas Health San Antonio’s Medical Arts and Research Neurology Clinic were screened using protocol-defined eligibility criteria: adults aged 40–75 years without a history of stroke, traumatic brain injury, seizures, or cognitive impairment. Nine ALS patients met eligibility criteria and were contacted by telephone to provide initial information and assess availability to attend a single 1–2-hour study visit at the clinic. Five patients provided consent to the study and completed study procedures. The protocol was approved by the University of Texas Health San Antonio Institutional Review Board (IRB Approval # STUDY00000628).

EEG data were collected from all participants (n = 5) using a low-cost, wireless EEG system with five electrodes positioned at AF3, AF4, T7, T8, and Pz (Emotiv Insight 2.0; EMOTIV Inc., San Francisco, CA, USA). Study procedures and data analysis were conducted using EmotivPRO^6^ software in conjunction with Python-based machine learning libraries, including Keras and TensorFlow.

### Word Selection and Subject Preparation

Participants completed an isolated-word task consisting of five words: “Yes,” “No,” “Help,” “Sun,” and “Water.” These words were selected for their relevance to basic caregiving needs and daily communication, encompassing both emotionally salient and neutral terms.

Prior to EEG acquisition, participants were first instructed to speak the words aloud in a fixed sequence (“Yes”, “No”, “Help”, “Sun”, and “Water”) to familiarize them with task timing. During EEG acquisition, auditory cues were delivered as beeps at one-second intervals to indicate the timing of each word, with each cue corresponding to an expected subvocalized word. To maintain temporal alignment, the word “Yes” was played audibly every 30 seconds (six cycles). Participants were instructed to alert the experimenter if they detected a sequencing error, at which point the recording was discarded and a new recording started.

Each EEG recording lasted for approximately 3 minutes. Participants completed multiple recordings as tolerated. Recordings were excluded if participants reported loss of focus or missed cues. EEG signal quality was continuously monitored and maintained above 83% in accordance with the manufacturer’s guidelines. Data from two participants were excluded due to movement-related artifacts, yielding analyzable datasets from three participants. A schematic overview of the study workflow is provided in **Supplemental Figure 1**.

### Data Collection and Preprocessing

EEG data were exported as comma-separated values (CSV) files. The original raw files contained timestamp metadata, task markers, and multichannel EEG signals from electrodes AF3, AF4, T7, T8, and Pz. Using Python and the Pandas library, datasets were reduced to the task label column and electrode-specific signals. Task labels corresponding to the subvocalized words were converted from string identifiers to numerical class labels using one-hot encoding.

For each included participant, recordings were partitioned into training, validation, and testing datasets to emulate a real-world deployment scenario in which an individual with ALS first calibrates the system prior to active use. Dataset composition was as follows: Participant #003 (3 training, 2 validation, 1 testing recordings), #004 (4 training, 2 validation, 1 testing recordings), and Participant #005 (5 training, 2 validation, 2 testing recordings). Data from Participants #001 and #002 were excluded due to excessive movement-related artifacts and inability to maintain task engagement.

All EEG signals were filtered using a Butterworth bandpass filter (0.3-60 Hz) with sampling frequency 128 Hz, corresponding to the acquisition rate of the Emotiv Insight 2.0. A 60 Hz notch filter with a quality factor of 30.0 was applied to attenuate line noise.

Filtered signals were reformatted into image-like tensors, with each image sample represented as a 128 x 5 single-channel array corresponding to one second of EEG data across five electrodes. Resulting datasets `(x_train, x_val,` and `x_test)` comprised three-dimensional arrays of stacked samples, with corresponding encoded labels stored in `y_train`, `y_val`, and `y_test,` respectively. For example, if the fifth word in the training dataset was “Water,” `x_train[4]` (the fifth element in that array) will be the 128 x 5 array containing the data for thinking the word “Water” and `y_train[4]` will return `[0. 0. 0. 0. 1.]`, denoting the word “Water.” All preprocessing and data-handling routines were implemented as modular functions to enable real-time execution in future system extensions.

### Model Architecture and Training

Artificial neural networks are computational models inspired by the organization of biological neural systems and consist of interconnected nodes that learn mappings between input data and output labels. Convolutional neural networks (CNNs) extend this framework by incorporating convolutional layers that enable efficient extraction of structured patterns, making them well suited for signal- and image-based classification tasks.

Multiple CNN models were trained using subject-specific datasets, with each model trained exclusively on data from a single participant. All models were implemented as one-dimensional (1D) CNNs using Keras and Tensorflow. The best-performing architecture, using notation from Wang et al.,^7^ consisted of sequential convolutional, pooling, normalization, and fully connected layers with scaled exponential linear unit (SELU) activations and dropout-based regularization:

C25@11×2 - SELU - Dropout(0.5) - BN - C25@11×1 - SELU - P5×3 - C50@11×1 - SELU - SpatialDropout(0.5) - BN - P5×3 - C100@11×1 - SELU - SpatialDropout(0.5) - BN - P3×3 - C100@11×1 - SELU - P2×2 - Flatten - F5 (Softmax)

Models were trained using the Adam (Adaptive Moment Estimation) optimizer with a learning rate of 0.001, a batch size of 128, and a maximum of 100 epochs. Proportional class weighting was applied to address class imbalance and improve classification performance.

### Data analysis

Accuracy, precision, recall, and F1-scores were computed, and statistical significance was evaluated using binomial testing. For comparative evaluation, model performance was also assessed against a support vector machine (SVM) and deep neural network (DNN). The SVM employed a sigmoid kernel and represented a non-neural-network-based statistical classifier previously applied to EEG data, whereas the DNN shared a fully connected architecture with the CNN, but lacked convolutional layers. DNN’s hyperparameters were optimized using Hyperband, an efficient alternative to Bayesian optimization.

## Results

Three participants were included in the final analysis; all were male, aged 56 to 67 years, right-handed, cognitively intact, and disease duration between 1 to 5 years. Revised ALS Functional Rating Scale (ALSFRS-R) scores of the participants ranged from 25 to 40.

A total of 3,819 EEG recordings were analyzed, each corresponding to a 1-second EEG recording segment of a subvocalized word. **Figure 1** presents boxplots of classification accuracy across five independent training runs per study participant.

**Figure 1.**
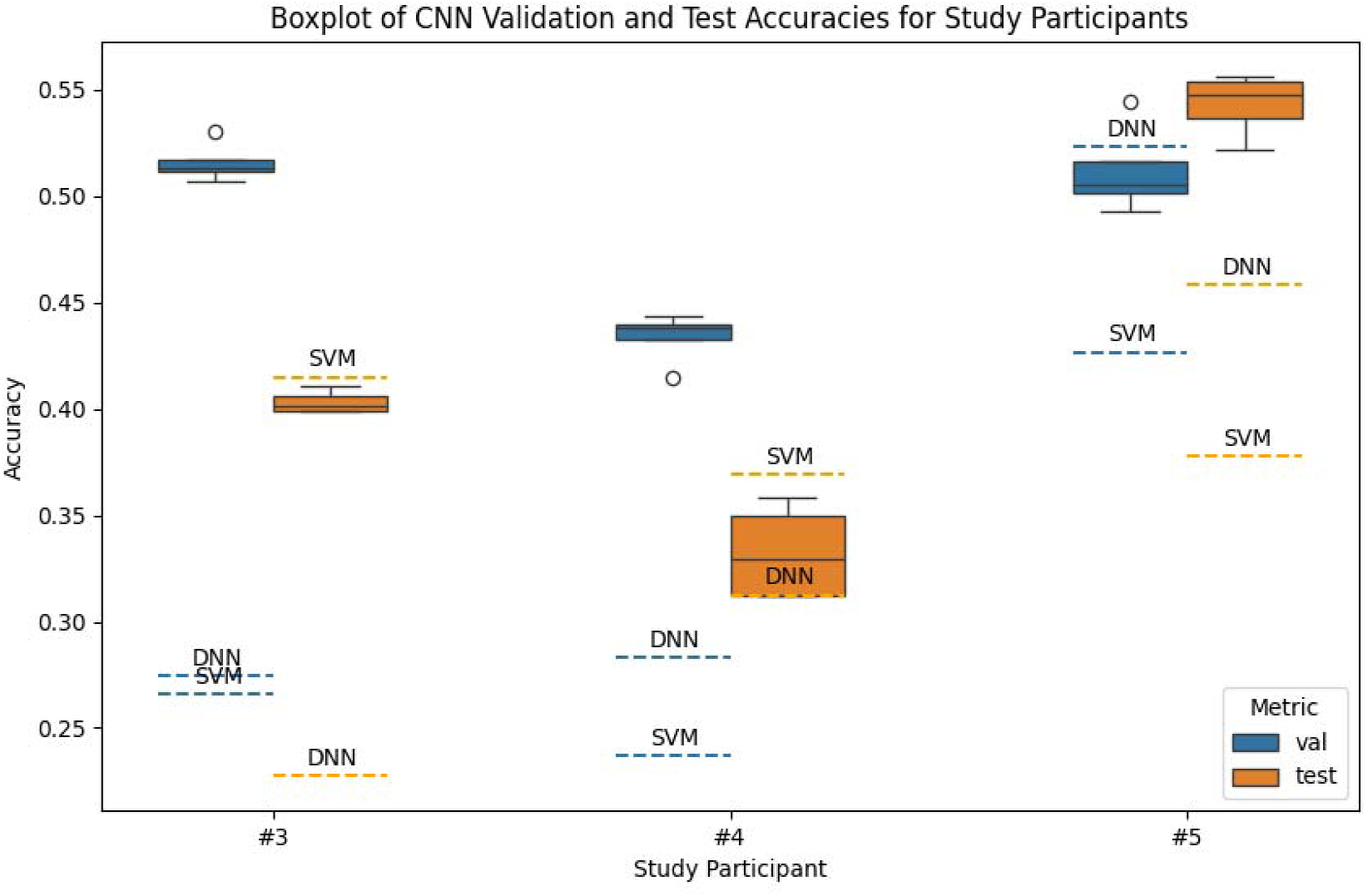
Boxplots of validation and test classification accuracies across five independent training trials.

Median test accuracies across five trials for the CNN models of three study participants were 40.17%, 32.95%, and 54.76%, respectively – all significantly exceeding chance-level performance (20%, p <0.001). Learning curves during training and validation are presented in **Supplemental Figures 2-4**. **Figure 2** shows the confusion matrices for each study participant derived from the testing dataset following a single model run, comparing predicted class labels with actual subvocalized words. Confusion matrices from the validation datasets of three participants are presented in **Supplemental Figures 5-7**. For example, as illustrated in **Figure 2a**, the model correctly classified 23 instances of the word “Yes” in the testing dataset.

**Figure 2.**
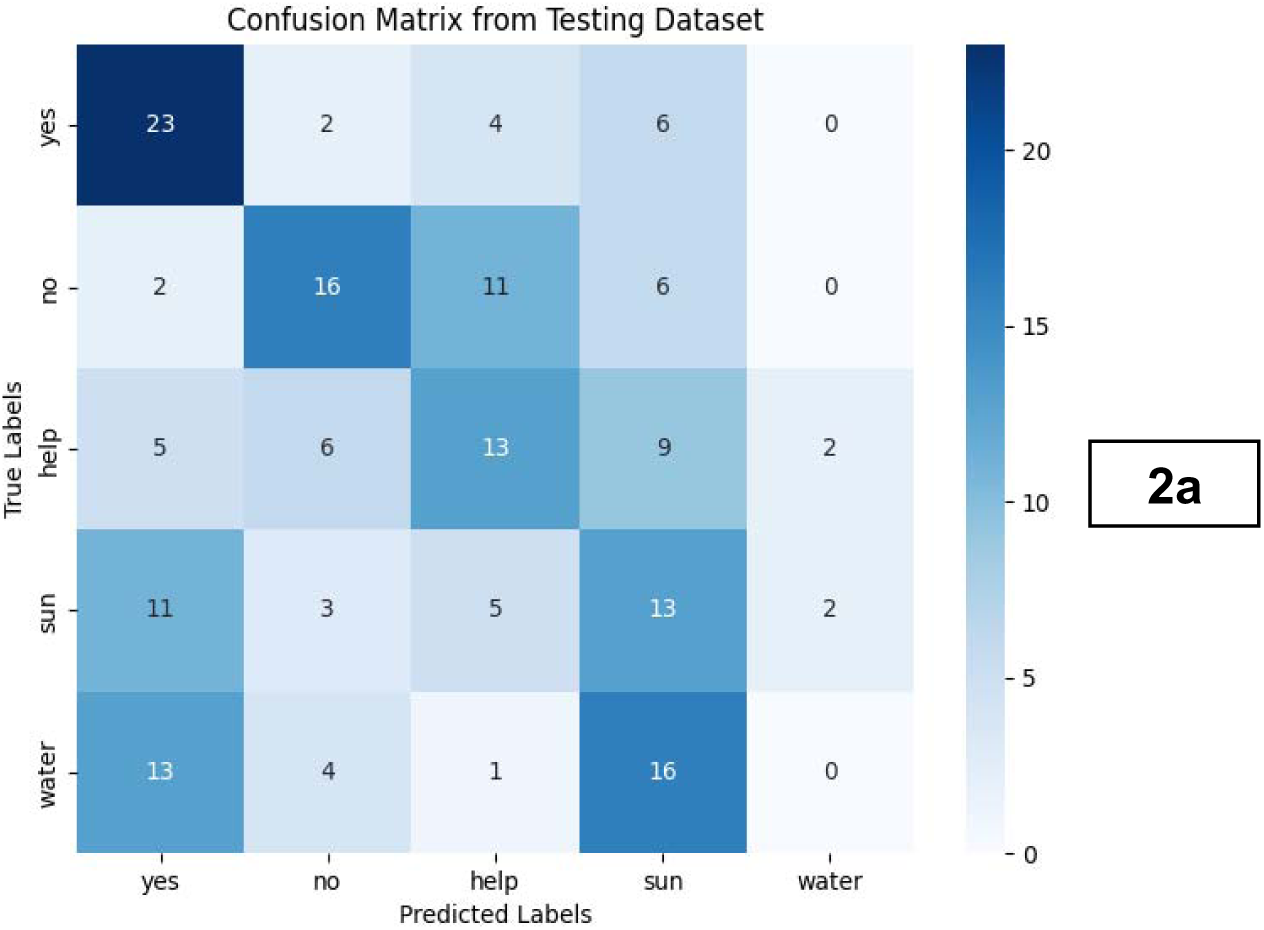
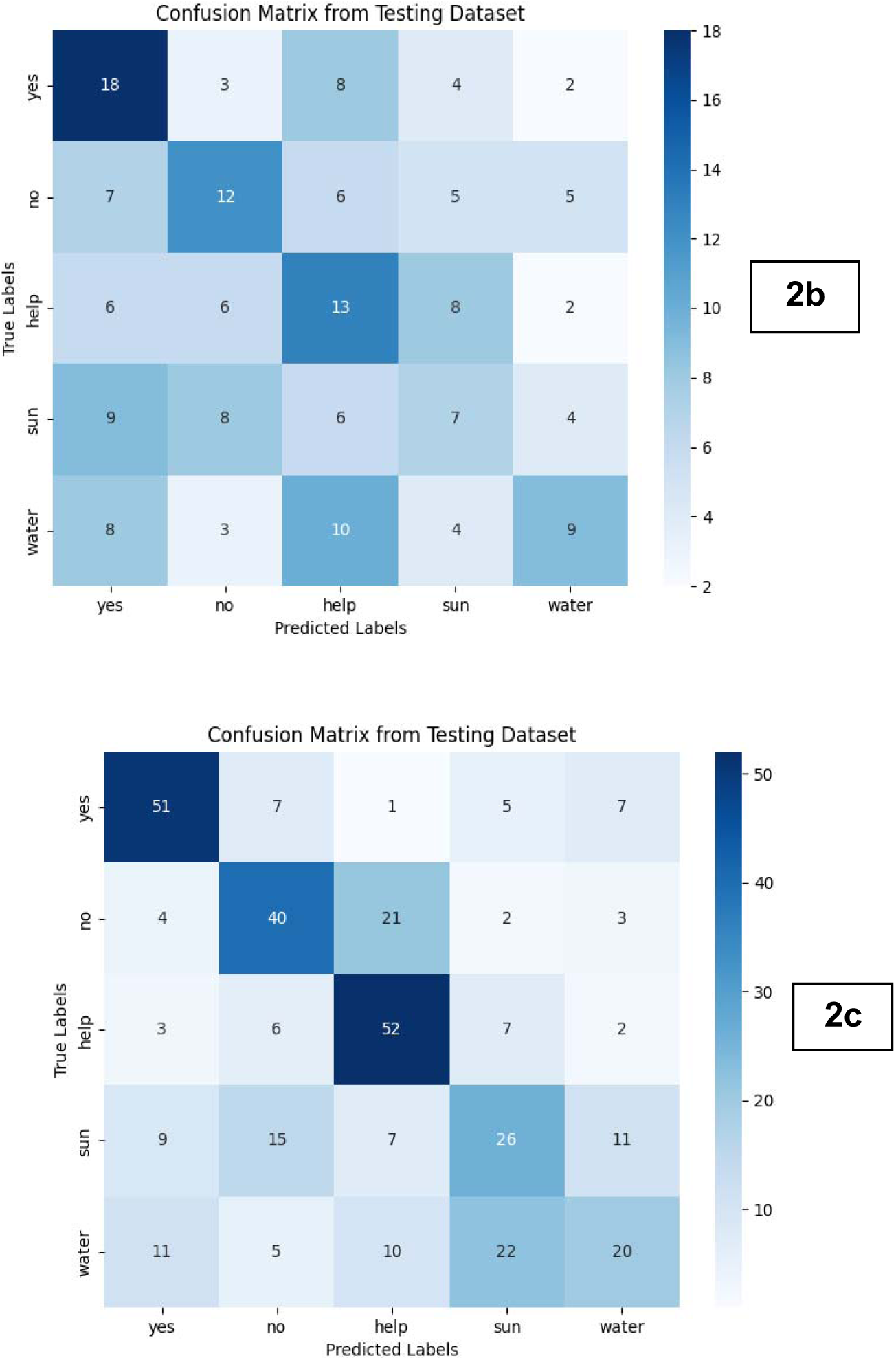
Confusion matrices derived from the testing dataset for each study participant: **(2a)** Participant #003 (top), **(2b)** Participant #004 (middle), and **(2c)** Participant #005 (bottom).

## Discussion

This proof-of-concept study demonstrates that EEG signals associated with subvocalized word production can be detected and decoded in individuals with ALS using a low-cost, noninvasive BCI coupled with a deep learning model. By targeting cortical activity related to speech planning rather than overt motor execution, this approach supports a potential communication modality that bypasses the degenerating corticobulbar pathways that underlie anarthria in advanced ALS.

Model performance was highest in Participant #005, despite this individual having a total ALSFRS-R score of 25 and a speech sub score of 0, consistent with complete loss of functional speech (anarthria). This outcome was anticipated, as this participant contributed the largest dataset and exhibited fewer motion-related artifacts, aided by the use of a neck brace to stabilize upper trunk weakness. In contrast, Participants #003 and #004 – both of whom retained normal speech function (ALSFRS-R speech score = 4) – demonstrated higher performance during validation than testing overfitting in the setting of limited training data and known inter-trial variability in EEG signals. Notably, model performance remained statistically significant across participants with a wide range of ALSFRS-R (all p <0.001), suggesting that this approach may generalize across disease severity. Overall, CNN consistently outperformed both the SVM and DNN comparators in terms of accuracy and stability.

Words with greater emotional or communicative salience (“Yes,” “No,” and “Help”) were decoded more accurately than with more neutral nouns such as “Sun” and “Water,” consistent with prior evidence that affectively meaningful stimuli elicit more stereotyped and discriminable neural responses.^8^

Several limitations warrant consideration. Data collection was restricted to a single session per participant due to clinical scheduling and time constraints. EEG signals are inherently susceptible to noise from attention, fatigue, and movement. Additionally, non-cognitive artifacts may confound neural classification. For instance, consistent motor behaviors (such as deep breathing) while a subject subvocalizes a specific word may cause the model to recognize that artifact rather than the intended neural representations. Nonetheless, these limitations reflect real-world conditions and underscore the need for artifact-robust algorithms and longitudinal refinement, which may enable individuals with severe expressive communication impairment to interact with caregivers, family members, and everyday technologies in home-based settings.

Despite these limitations, this study provides initial evidence that a noninvasive, low-cost EEG-based BCI technology integrated with a deep learning model can decode subvocalized words in ALS patients. Unlike existing invasive and resource-intensive BCI systems, the approach described here is portable, affordable, and comparatively simple to deploy, supporting its potential accessibility across socioeconomic contexts. With further validation and longitudinal refinement, such systems may enable individuals with severe expressive communication impairment to interact with caregivers, family members, and everyday technologies in home-based settings.

## Conclusion

Using a low-cost, noninvasive EEG system, the proposed CNN model achieved statistically significant decoding of subvocalized words in individuals with ALS, with accuracies up to 54.76%, exceeding the expected chance-level performance of 20% under the null hypothesis. Although current accuracy levels are insufficient for reliable real-world communication, they establish a promising foundation for future methodological refinement. Future studies incorporating larger cohorts, repeated recording sessions, and longitudinal training are warranted to improve decoding performance and advance noninvasive neural speech decoding toward practical application for individuals with severe expressive communication impairment.

## Data Availability

All data produced in the present study are available upon reasonable request to the authors.

## Acknowledgements

The authors are deeply grateful to the participants without whom this proof-of-concept study would not have been possible.

## Disclosure

The authors report no relevant disclosures.

## Funding

The authors report no targeted funding.

## Data Sharing Statement

Deidentified data that support the findings of this study may be made available from the corresponding author upon reasonable request.

## Supplemental Figures

**Supplemental Figure 1.**
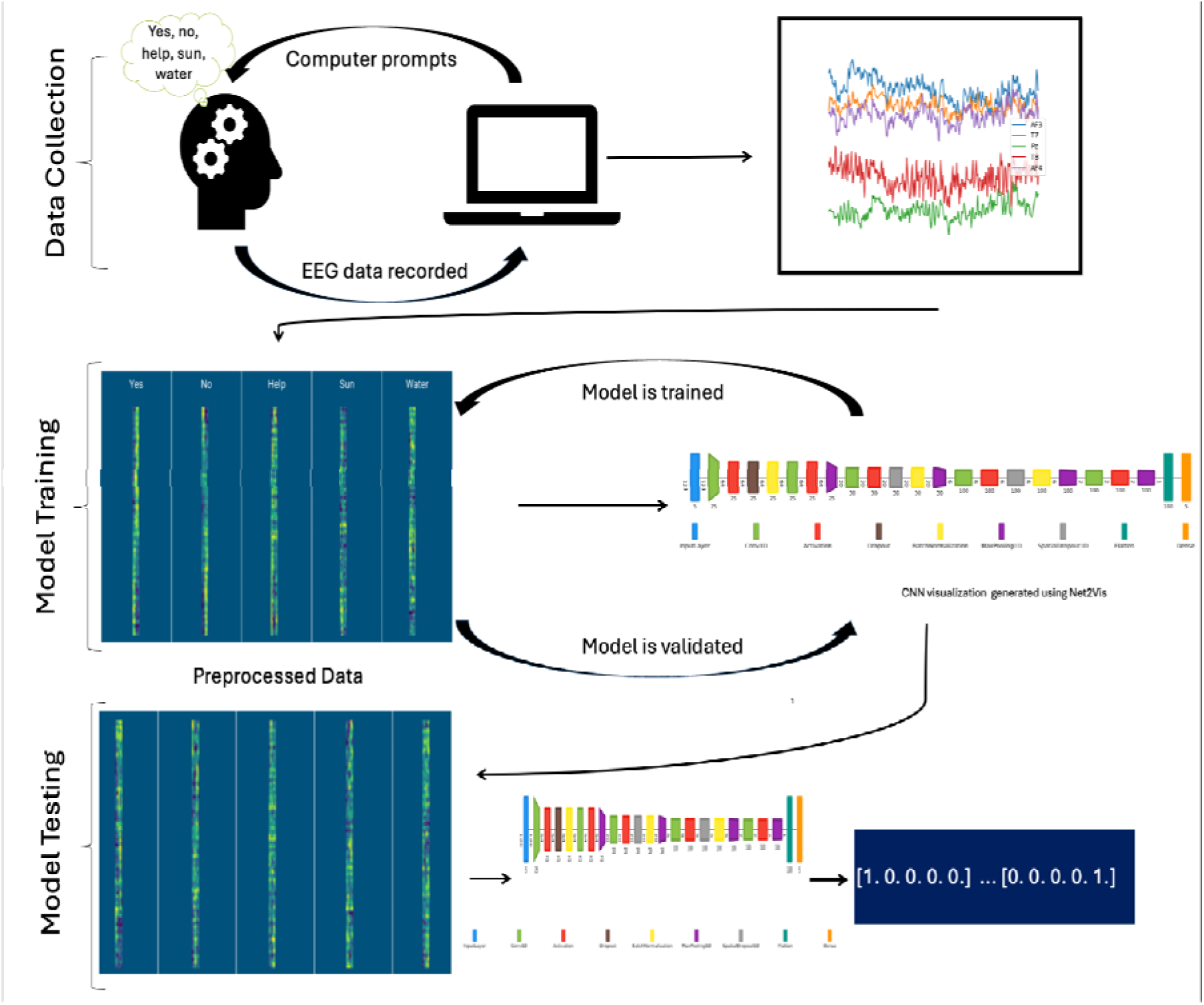
Schematic overview of the study workflow.

**Supplemental Figure 2:**
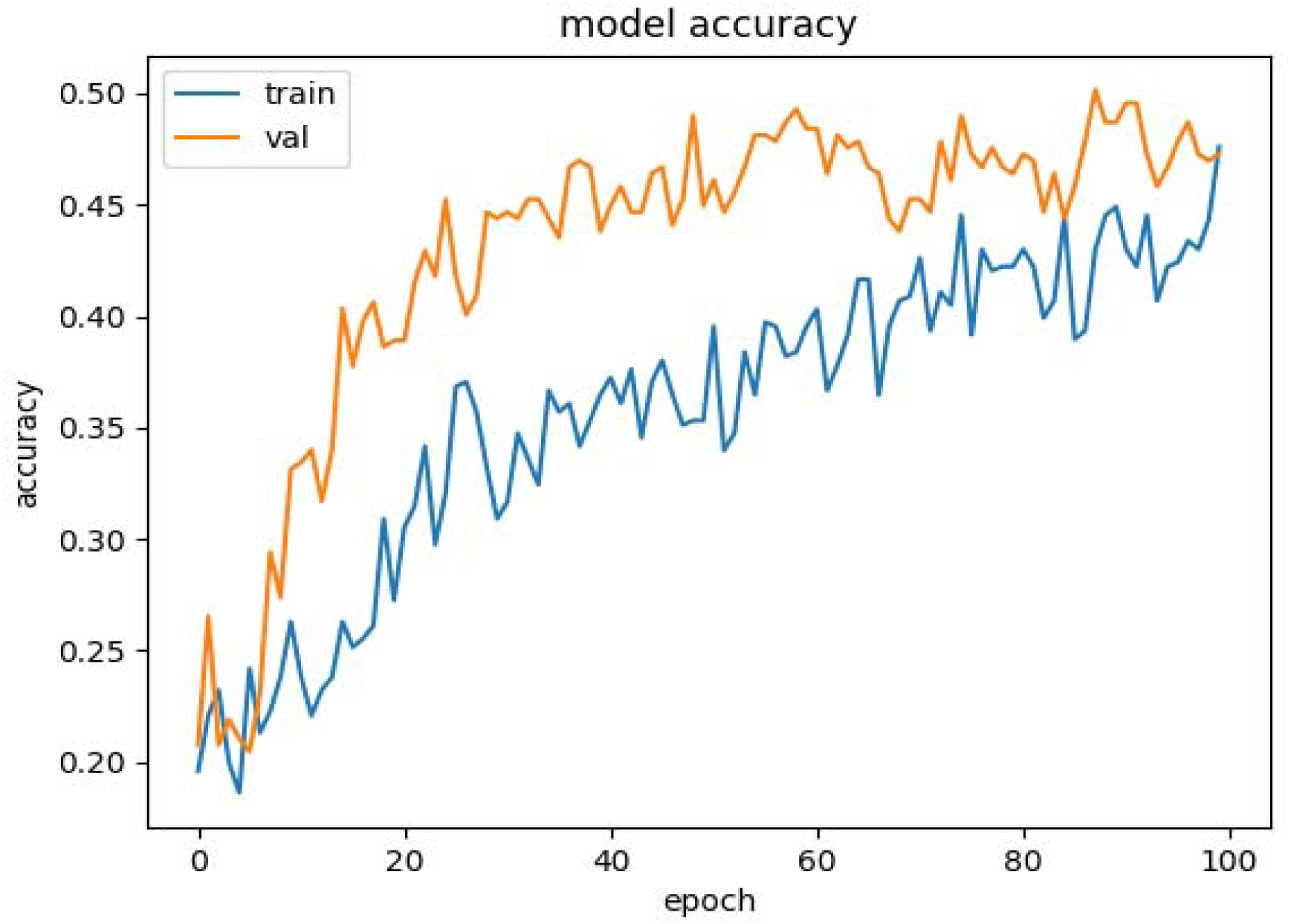
Model accuracy over epochs for study participant #3.

**Supplemental Figure 3:**
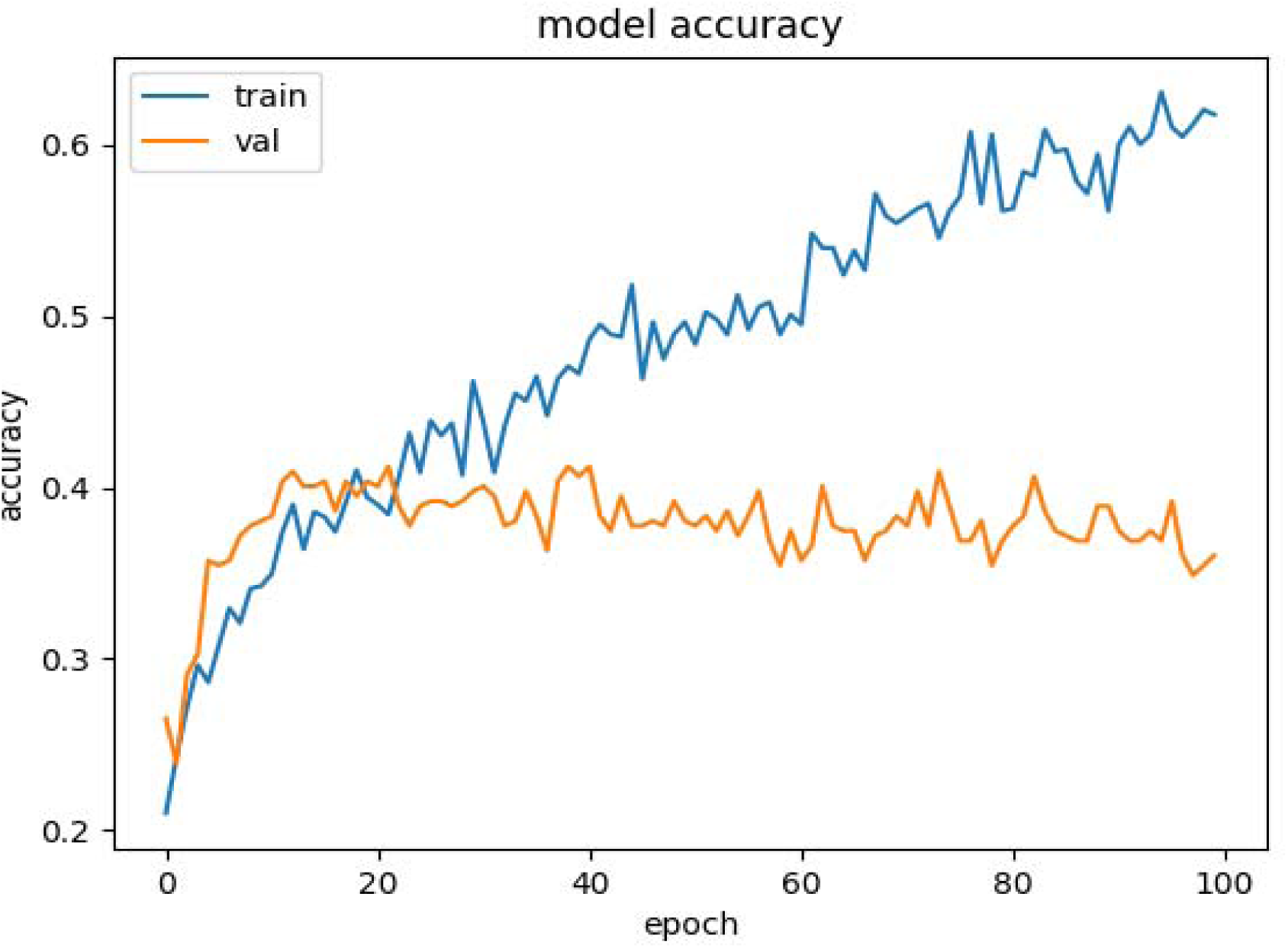
Model accuracy over epochs for study participant #4.

**Supplemental Figure 4:**
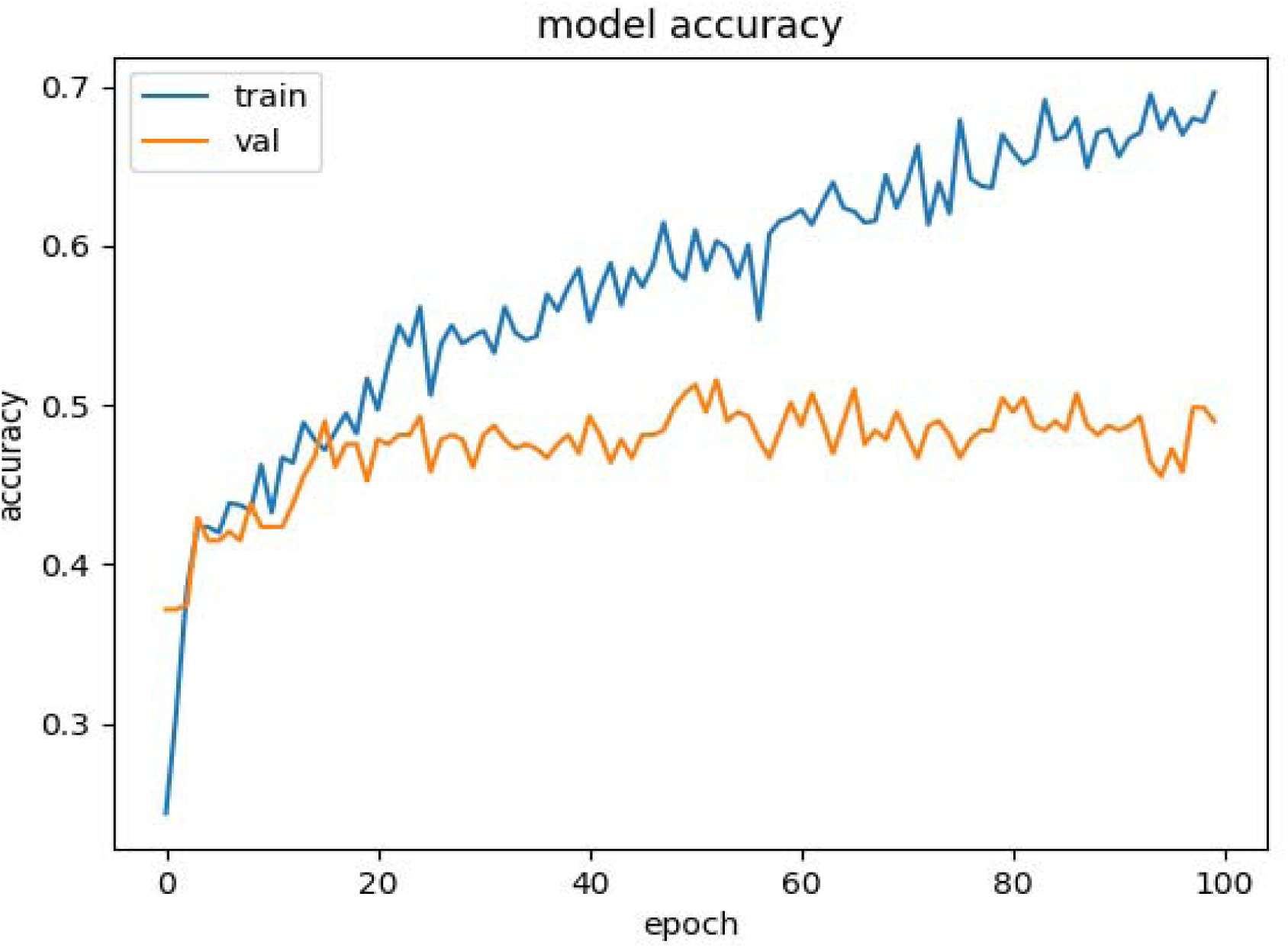
Model accuracy over epochs for study participant #5.

**Supplemental Figure 5:**
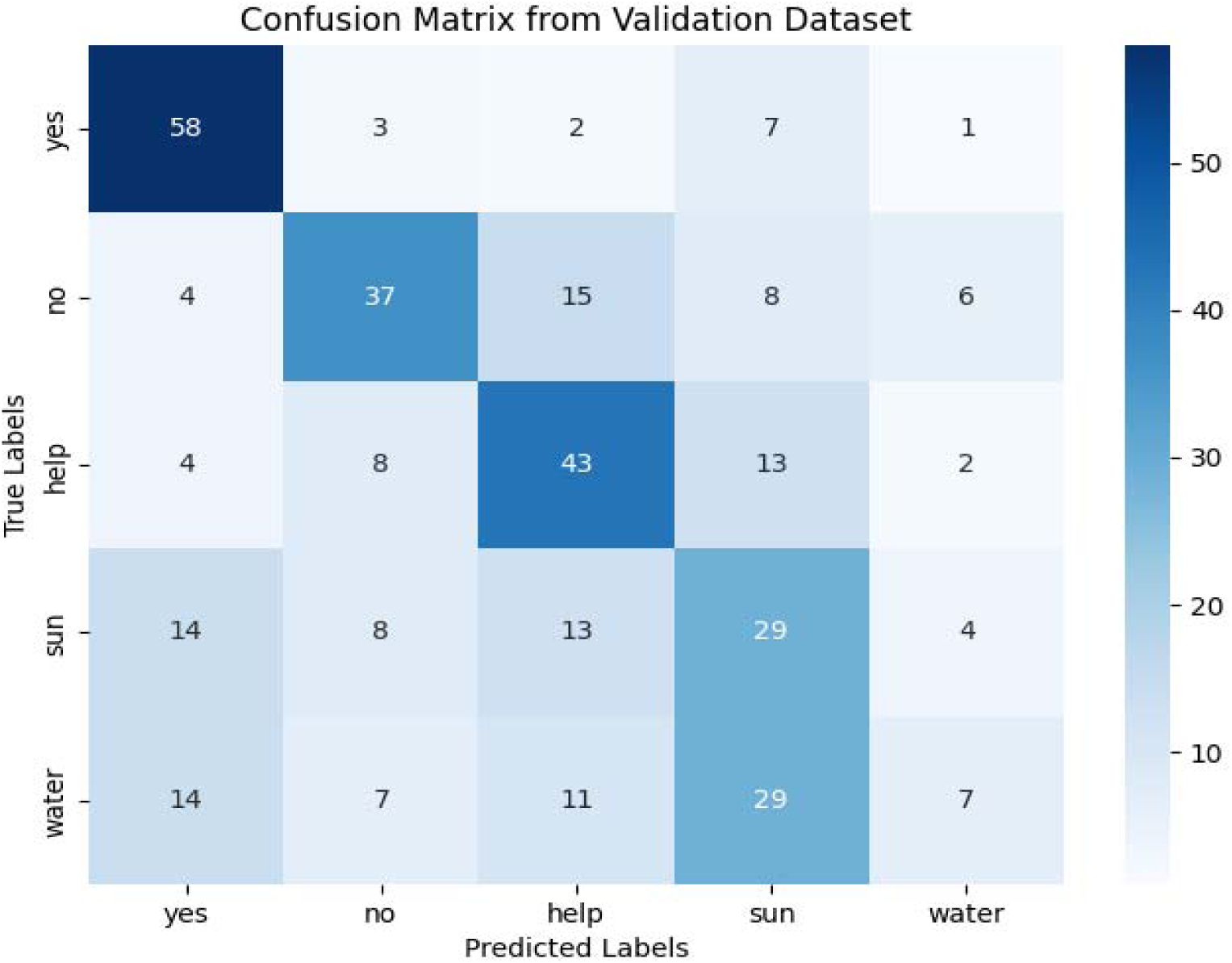
Confusion matrix from validation dataset for study participant #3.

**Supplemental Figure 6:**
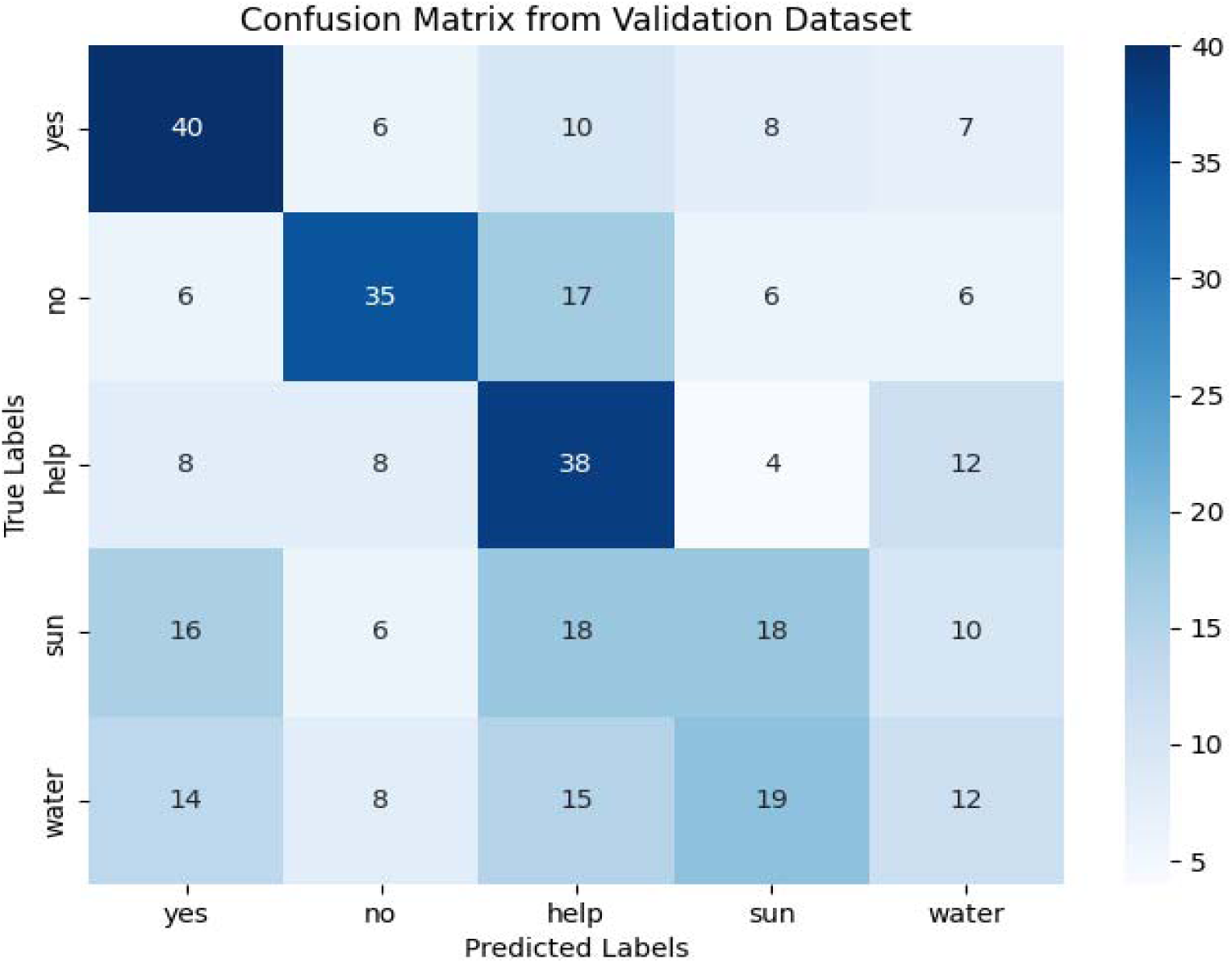
Confusion matrix from validation dataset for study participant #4.

**Supplemental Figure 7:**
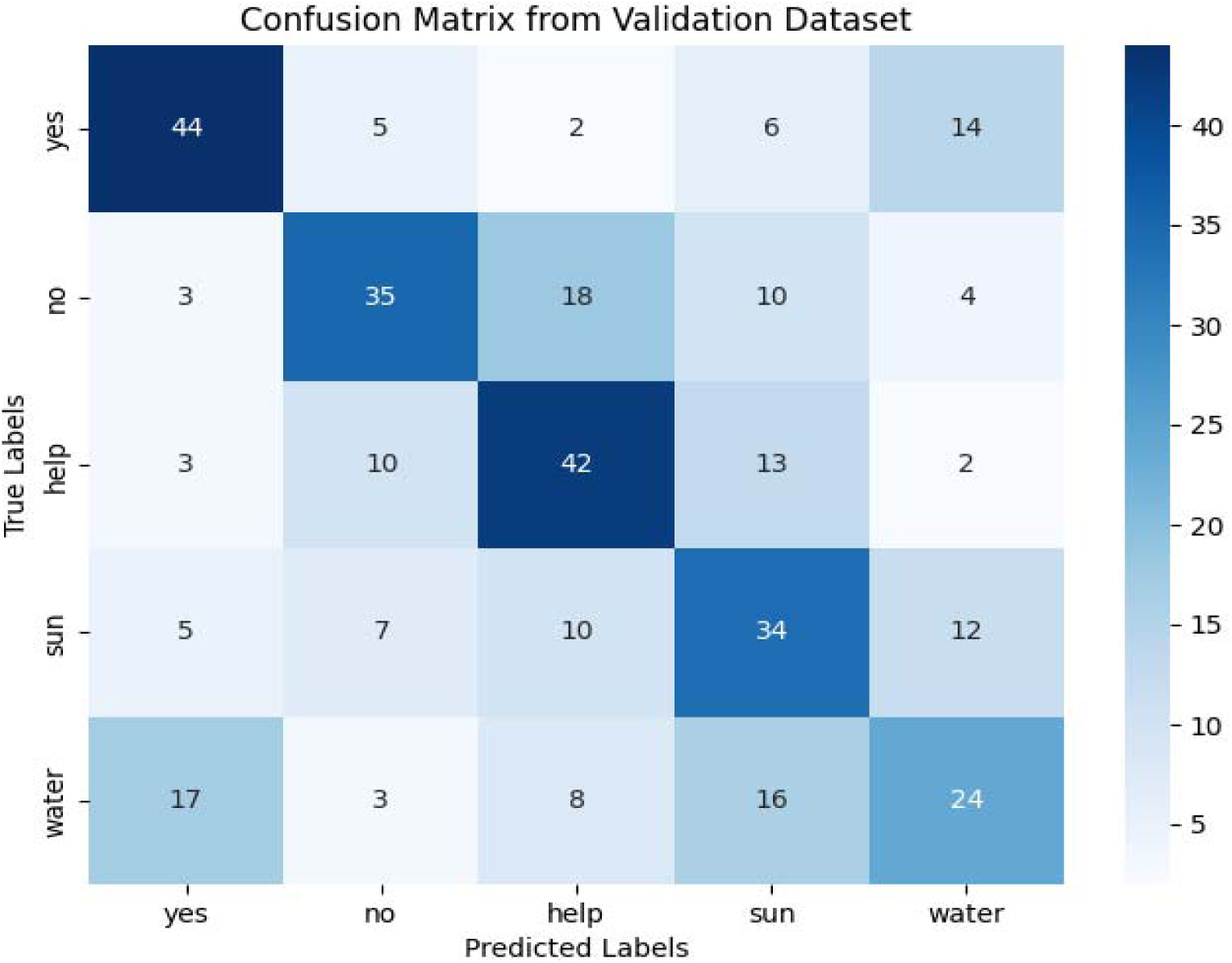
Confusion matrix from validation dataset for study participant #5.

## Notes

### Competing Interest Statement

The authors have declared no competing interest.

### Funding Statement

This study did not receive any funding.

### Author Declarations

University of Texas Health San Antonio Institutional Review Board (IRB)

### Summary of Updates

In Results section: Sentence "Learning curves during training and validation are presented in Supplemental Figures 1-3." should read as "Learning curves during training and validation are presented in Supplemental Figures 2-4." In Results section: Sentence "Confusion matrices from the validation datasets of three participants are presented in Supplemental Figures 4-6." should read as "Confusion matrices from the validation datasets of three participants are presented in Supplemental Figures 5-7." In Results section: Figure 2 was mislabeled with 3a, 3b, 3c. Revised as 2a, 2b, 2c.

